# The connection between COVID-19 vaccine abundance, vaccination coverage, and public trust in government across the globe

**DOI:** 10.1101/2022.05.24.22275504

**Authors:** Ida G. Monfared

## Abstract

This study investigates that how the number of COVID-19 vaccines secured correlates with the vaccination coverage (full and booster) depending on whether there is trust in national government or not across 47 countries. The data are based on global figures as of Nov. 2021 and Feb. 2022 while measures for confidence in government is according to Gallup World Poll, Oct. 2021. The model includes an interaction term of the two key variables, also controls for a range of socio-economic factors and country specific variables. The results indicate a non-linear and mixed relationship between the number secured, the public trust, and the vaccination rate. In Feb. 2022, with confidence in government, securing number of vaccines to cover 200% of the population (or more) increased the full vaccination rate by 12.26% (95% CI: 11.70 - 12.81); where number secured was 300% (or more), the coverage increased by 7.46% (95% CI: 6.95 - 7.97). Under similar scenarios, rate of booster shots increased by 13.16% (95% CI: 12.62 - 13.70; *p* < 0.01) and 14.36% (95% CI: 13.86 - 14.85; *p* < 0.01), respectively. Where the number secured fell below 200%, confidence in government had a revers relationship with the rate of full vaccination (-2.65; 95% CI: -3.32 - -1.99), yet positive with the rate of booster shots (1.65; 95% CI: 1.18 - 2.12). These results indicate that better success can be achieved by a combination of factors including securing sufficient number of vaccines and also ensuring the public trust. Vaccine abundance, however, cannot be translated into greater success in vaccination coverage. This study highlights the importance of efficiency in acquiring vaccine resources and need for improvement in public belief in immunization programmes rather than stock piling.

## Introduction

The rapid development of Covid-19 vaccine and its rollout have set an unprecedented record. Since the approval of the Pfizer/BioNTech vaccine by the World Health Organisation (WHO) in December 2020, approximately 11.3 billion doses of vaccines have been administered[1] within 28 months. There has been, however, a staggering imbalance in getting access to the vaccine supplies across the world. Some countries, often with higher income, placed early orders of vaccines in volumes that were multiple times the number of their population[2] and after the provision of two-dose vaccinations are now in the process of rolling out booster shots. Meanwhile, some other countries, often low-income, are yet to vaccinate their medical staff and vulnerable populations, even with a single dose[3]. Naturally, during times of crises, it is understandable that policymakers prioritize their own populations. The question remains, however, whether securing abundant resources necessarily translates into a better vaccine coverage of the population when social factors and, in particular, the public’s confidence in national government is considered.

Confidence in government (CiG) is an emerging common predictor that can influence the public’s willingness to be vaccinated [4–7]. Despite the heterogeneity of countries in terms of cultural norms and setting, CiG has not only been detected in the studies of high-income countries (HICs), but also was found to be an important contributor in studies of low and middle-income countries (LMICs), with higher degrees of mistrust in government being linked to the vaccine hesitancy [8–11].

Potential harm from the under and overuse of medical resources and global disparities have been noted long before the recent pandemic[12]. In relation to the COVID-19 vaccines, there is growing evidence of unused and hard to redistribute stock piles in HICs were the demand has dropped[13,14]. This is not only a waste of life-saving medical resources but also funds for HICs - both of which could perhaps be placed in better use elsewhere.

The present study investigates to what extent the combination of these factors and, in particular, the number of vaccine resources secured at the procurement stage, and CiG at the recipient end, might influence the rate of full vaccination and booster coverage across the world.

## Method

In this study, the figures for CiG are obtained from the Gallup World Poll, October 2021 and the question WP139:*“Do you have confidence in the national government?*” with binary response options “*Yes*” or “*No*[15], treating “Don’t know” and no responses as missing. Although they are not specifically asked in relation to COVID-19 vaccines, these figures indicate the degree to which the public trust the national government and, subsequently, its actions and policies. The data on the number of vaccines secured adjusted for country population in percentage is based on figures provided by the International Monetary Fund (IMF)[16] and vaccine coverage figures (full vaccination per 100 people and booster rates) are based on the data reported by Our World in Data[1]. For the present study, the main focus was placed on two snapshots of these data as of November 2021 and February 2022 (the last day of the month when the data was available). These dates were chosen as November was the closest date to the Gallup poll data, considering any potential lags in vaccines uptake, and February 2022 by when, in many countries, both full vaccination and booster jab programmes were well in place. Figure 1 illustrates the number of vaccines secured and the ratio of the population that has been fully vaccinated as of February 2022. In this figure, those countries where CiG was lower than 50% are shown in triangles and those above 50% are shown in dots (please see Figure A1 in Appendix A that depicts a picture of larger number of countries without the inclusion of the Gallup data).

**Figure 1.**
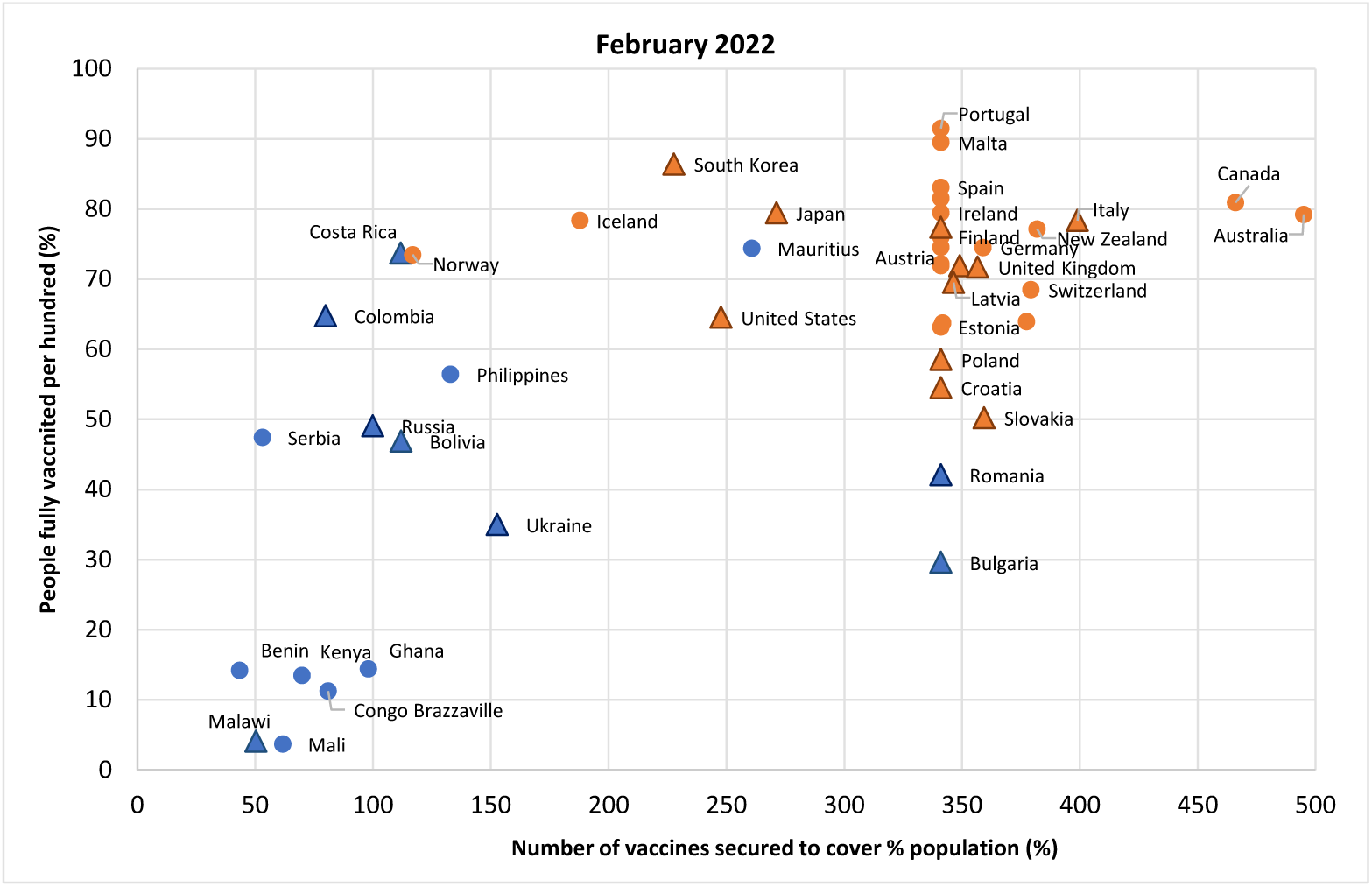
The number of vaccines secured and the ratio of population that were fully vaccinated as of February 2022. Countries coloured in blue indicate LMICs and orange indicate HICs. Triangles are countries where below 50% of the poll surveys said they have CiG and dots are those where more than 50% said they have CiG.

For the numbers secured, two set of thresholds are noticeable; a 200% threshold that could suffice full vaccination of the population (assuming the two-dose requirement), and a 300% threshold that exceeds the full vaccination needs, yet could secure the numbers required for providing booster shots. Several European countries had the same number of vaccines secured as they acted as a united entity. In order to identify the efficacy of the number of vaccines secured, these two thresholds were used and defined as dummy variables below which countries were coded as 0, and above as 1. This approach allowed us to investigate whether securing more vaccine supplies improve the rate of vaccination coverage when CiG is also considered.

Equation (1) presents the study model that is a multiple regression including an interaction term of number secured and CiG[17]:

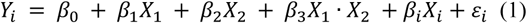

where:

*Y*_*i*_ : % of population fully vaccinated per 100 people / received booster shots;

*β*_1_: coefficient for having CiG when the number of vaccines secured is below the dummy threshold;

*β*_2_: coefficient for the number of vaccines secured when the there is no confidence in the national government (CiG = 0);

*β*_3_: the interaction term between CiG and the dummies for number of vaccines secured;

*β*_*i*_: coefficients for the covariates included in the model (gender, age, employment, per capital person income quantile, marital status, health status, access to internet, being born in the country, education level, living in urban / rural areas, country population density, GDP per capita;

*ε*_*i*_: error term.

Here, robust standard errors were used to address potential heteroskedasticity and throughout, the sample weight was applied. Stata 16.1 was used for the analysis.

## Results

The final dataset covers 47 countries with *N* = 47,656 observations. Descriptive figures for pooled data are presented in Table 1 and other key variables for each country are presented in Appendix A, Table A1.

**Table 1.** Characteristics of respondents across all 47 countries *N* = 47,656 (weighted).

| Characteristic |  | (% / mean (sd.)) |
| --- | --- | --- |
| Gender | Male | 48.44 |
| Age |  | 44.87 (18.78) |
| Employed (any type of employment) | Yes | 60.26 |
| Per capita income quintiles | Poorest 20% | 19.9 |
|  | Second 20% | 20.02 |
|  | Middle 20% | 19.99 |
|  | Fourth 20% | 19.99 |
|  | Richest 20% | 20.09 |
| Married / Domestic partner | Yes | 53.8 |
| Health problems | Yes | 24.34 |
| Access to internet | Yes | 83.74 |
| Born in the country | Yes | 90.69 |
| Education level | Completed elementary education or less. | 21.84 |
|  | Secondary, 3-year tertiary secondary education and some education beyond secondary education. | 58.1 |
|  | Completed four years of education beyond high school and/or received a 4 - year college degree. | 20.06 |
| Living in rural / urban area | A rural area or on a farm | 17.19 |
|  | A small town or village | 39.08 |
|  | A large city | 28.82 |
|  | A suburb of a large city | 14.9 |

Table 2 presents the outcome of running equation (1) for Nov. 2021, comparing the results for when the number of vaccines secured is not included (column 2) with when thresholds of 200% (column 3) and 300% are included (column 4). Column 1 illustrates the difference that ex/inclusion of country GDP per capita makes before incorporating the interaction term (the complete list of covariates coefficients is presented in Appendix A, Tables A2).

**Table 2.** The coefficient of association between the rate of people fully vaccinated per hundred in Nov. 2021 and the interaction between CiG, and number of vaccines secured for population by thresholds of 200% and 300%.

|  | Number secured<br>and GDP<br>excluded | Number secured<br>excluded | Secured 200%<br>or more | Secured 300%<br>or more |
| --- | --- | --- | --- | --- |
|  | (1) | (2) | (3) | (4) |
| CiG = 1 | 1.89***<br>(1.43 - 2.35) | -1.70***<br>(-2.01 - -1.39) | -2.69***<br>(-3.26 - -2.12) | -3.61***<br>(-4.08 - -3.14) |
| Secured 200% or more = 1 |  |  | 12.33***<br>(11.80 - 12.86) |  |
| CiG # Secured 200% or more |  |  | 3.36***<br>(2.71 - 4.01) |  |
| Secured 300% or more = 1 |  |  |  | 6.84***<br>(6.42 - 7.26) |
| CiG # Secured 300% or more |  |  |  | 4.50***<br>(3.91 - 5.10) |
| GDP per capita |  | 0.00***<br>(0.00 - 0.00) | 0.00***<br>(0.00 - 0.00) | 0.00***<br>(0.00 - 0.00) |
| Constant | 16.02***<br>(14.54 - 17.50) | 8.99***<br>(7.97 - 10.01) | 11.44***<br>(10.44 - 12.43) | 10.93***<br>(9.94 - 11.92) |
| Observations | 46,819 | 46,819 | 46,819 | 46,819 |
| R-squared | 0.35 | 0.70 | 0.74 | 0.73 |

Robust 95% CI in parentheses; \*\*\* p<0.01, \*\* p<0.05, \* p<0.1.

Without including GDP and the number of vaccines secured into the model, the rate of population fully vaccinated per hundred has a positive and significant correlation with CiG. However, when countries GDP per capita is controlled for, this association becomes reverse. For CiG = 0, the rate of vaccinated increases by 12.33% (95% CI: 11.80 - 12.86; *p* < 0.01) when numbers secured is sufficient for 200% of the population (or more). Yet, this rate increases only by 6.84% (95% CI: 6.42 - 7.26; *p* < 0.01) when the number secured is 300% (or more). When CiG =1, securing the number by 200% increases the rate of coverage by 15.69% (95% CI: 15.16 - 16.21; *p* < 0.01) while with securing 300% and more, the coverage would increase by 11.34% (95% CI: 10.89 - 11.79; *p* < 0.01). On the other hand, when the number secured is below 200%, having CiG has a reverse relationship with vaccine coverage, ceteris paribus. Here, if GDP is not controlled for, this relationship becomes positive but statistically insignificant. Where the number of vaccines secured is above 200%, having CiG increases the coverage by 0.66% (95% CI: 0.34 - 0.99; *p* < 0.01). When the vaccines are secured by 300% and more, CiG =1 increase the coverage by 0.89% (95% CI: .053 - 1.26; *p* < 0.01).

Table 3 presents the results of running the same model for February 2022 for the rate of full vaccination and also the booster shots per hundred people.

**Table 3.** The coefficient of association between the rate of people fully vaccinated / received booster shots per hundred in Feb. 2022 and the interaction between CiG, and number of vaccines secured for population by thresholds of 200% and 300%.

|  | Full vaccination |  |  |  | Booster shots |  |  |  |
| --- | --- | --- | --- | --- | --- | --- | --- | --- |
|  | Number secured and GDP excluded<br>(1) | Number secured excluded<br>(2) | Secured 200% or more<br>(3) | Secured 300% or more<br>(4) | Number secured and GDP excluded<br>(5) | Number secured excluded<br>(6) | Secured 200% or more<br>(7) | Secured 300% or more<br>(8) |
| CiG = 1 | 1.94***<br>(1.50 - 2.38) | -1.21***<br>(-1.53 - -0.89) | -2.65***<br>(-3.32 - -1.99) | -2.63***<br>(-3.19 - -2.06) | 4.10***<br>(3.64 - 4.55) | 0.44***<br>(0.12 - 0.77) | 1.65***<br>(1.18 - 2.12) | 2.11***<br>(1.63 - 2.58) |
| Secured 200% or more = 1 |  |  | 8.71***<br>(8.13 - 9.29) |  |  |  | 13.37***<br>(12.86 - 13.89) |  |
| CiG # Secured 200% or more |  |  | 3.55***<br>(2.81 - 4.29) |  |  |  | -0.22<br>(-0.80 - 0.37) |  |
| Secured 300% or more = 1 |  |  |  | 4.51***<br>(4.04 - 4.99) |  |  |  | 16.16***<br>(15.69 - 16.63) |
| CiG # Secured 300% or more |  |  |  | 2.95***<br>(2.28 - 3.62) |  |  |  | -1.80***<br>(-2.37 - -1.23) |
| GDP per capita |  | 0.00***<br>(0.00 - 0.00) | 0.00***<br>(0.00 - 0.00) | 0.00***<br>(0.00 - 0.00) |  | 0.00***<br>(0.00 - 0.00) | 0.00***<br>(0.00 - 0.00) | 0.00***<br>(0.00 - 0.00) |
| Constant | 24.26***<br>(22.81 - 25.71) | 18.09***<br>(17.00 - 19.18) | 20.29***<br>(19.18 - 21.40) | 19.25***<br>(18.14 - 20.36) | 13.65***<br>(12.12 - 15.19) | 1.24**<br>(0.19 - 2.28) | 2.27***<br>(1.29 - 3.25) | 1.34***<br>(0.44 - 2.24) |
| Observations | 46,819 | 46,819 | 46,819 | 46,819 | 40,973 | 40,973 | 40,973 | 40,973 |
| R-squared | 0.35 | 0.65 | 0.67 | 0.66 | 0.17 | 0.59 | 0.65 | 0.70 |

Robust 95% CI in parentheses; \*\*\* p<0.01, \*\* p<0.05, \* p<0.1.

In terms of the full vaccination rate, the direction and scale of coefficients are rather similar to those for Nov. 2021 (columns 1-4 in Tables 2 and 3). For the rate of booster shots, amongst countries that are included in the present study, Iceland had the highest rate as of Feb 2022 with 67.23%, and the lowest was in Ghana with 0.32% (the chart that depicts the rate of boosters per hundred people and the number of vaccines secured is presented in Appendix A, Figure A2.). Dissimilar to figures for full vaccination rates, the relationship between rate of boosters and CiG remains positive and significant even when countries’ GDP per capita is controlled for. In the absence of CiG, securing the number of vaccines by 200% (or more), increases the booster rate by 13.37% (95% CI: 12.86 - 13.89; *p* < 0.01) and when the number secured reaches 300% (or more), this rate increases by 16.16% (95% CI: 15.69 - 16.63; *p* < 0.01). When the number secured is below 200%, having CiG increases the booster rate by 1.65% (95% CI: 1.18 - 2.12; *p* < 0.01). When CiG = 1, securing the vaccines by 200% increases the rate of boosters by 13.16% (95% CI: 12.62 - 13.70; *p* < 0.01). Changing this threshold to 300% correlates with a 14.36% (95% CI: 13.86 - 14.85; *p* < 0.01) increase in the rate of booster shots. Figure 2 illustrates the margin plots for various scenarios of the interaction term and its relationship with the outcomes (full vaccination and booster rates) as of February 2022.

**Figure 2.**
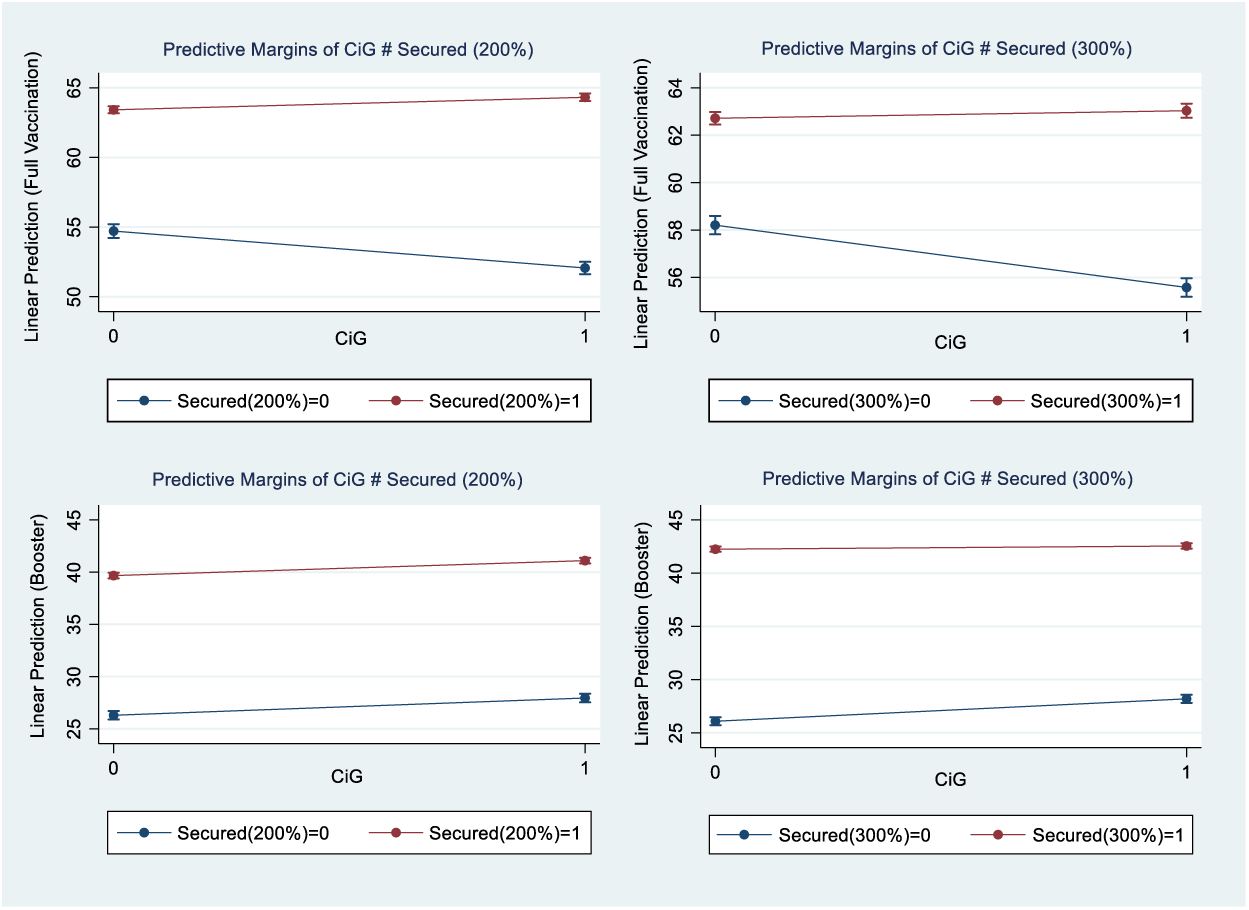
Interaction plot between CiG and number of vaccines secured for full vaccination/booster rate per hundred in Feb. 2022.

As can be seen, the interaction term of CiG and securing vaccines by 200% (or more) for booster shots is not significant (also see Table 3, column 7). Running a linear regression without an interaction term indicates the rate of boosters would increase by 13.27% (95% CI: 12.83 - 13.71; *p* < 0.01) if the number of vaccines secured is 200% (or more) and by 15.32% (95% CI: 14.93 - 15.71; *p* < 0.01) if the number secured is 300% (or more).

For robustness checks, I re-ran the analyses excluding Canada and Australia that are almost outliers with securing the numbers of vaccines sufficing to cover 466% and 495% of their population in February 2022, respectively, and it returned similar results (please see Appendix A, Table 5)^1^. It is also valid to argue for the role of workforce resources. For this, I tested the model by including the number of nurses per 10,000 people. This did not change the results substantially (Appendix A, Table A5). However, the number of nurses had a strong correlation with GDP per capita (*r* = 0.87; *p* < 0.01) and its inclusion could be an overfitting of the model, hence, it remained excluded.

## Discussion

Following the growing evidence that indicates the importance of CiG in vaccination uptake, the present study investigates how the interaction between this predictor and the number of vaccines secured correlates with the rate of full vaccination and also the booster shots across 47 countries.

The results indicate that the association between number of vaccines secured and rate of full vaccination varies for different scenarios, in presence or absence of CiG. Where there is no confidence in government, securing numbers of vaccines by 200% (or more) had a strong correlation with full vaccination rate, yet, when this number reaches to 300% (or more) it had a smaller impact. Where there was confidence in government, the rate of full vaccination increased even further by 3.36% (95% CI: 2.71-4.01) and 4.5%. (95% CI: 3.91 – 5.10), when the numbers secured were 200% and 300%, respectively.

On the contrary, when number of vaccines secured fell below 200%, CiG had a reverse relationship with the full vaccination rate. When this number was above 200%, having CiG increased the vaccination rate to a small degree and not substantially further more when the number secured reached 300% (or more). The directions of these relationships were similar in Nov. 2021 and Feb. 2022, with slightly smaller coefficients for the later date.

High inoculation rates, despite low CiG in some LMICs, e.g. Costa Rica and Colombia, can perhaps be explained by the strength of their vaccination campaigns and receiving vaccine supplies earlier than many other countries[18,19]. Moreover, primary evidence suggests that the potential for the acceptance of Covid-19 vaccines in these countries was particularly high and probably had roots in their longstanding belief in immunization programmes[20,21] that could go beyond the public’s confidence in present government. An imbedded belief in immunization programmes has also been reported as a contributor in the success of vaccination campaign in Portugal[22] and its relative failure in the US[23,24].

In relation to booster shots, the interaction term does not seem to be significant any longer, and higher rates of coverage are strongly correlated with the numbers secured reaching 200% or more, yet securing number by 300% or more increases this coefficient to a small degree. Here, CiG has a positive correlation with the rate of booster shots but is much smaller when compared to the effect from the numbers secured.

Undoubtedly, CiG plays a critical role in increasing the public’s acceptance of vaccines[25]. The present study, however, indicates that this association is not linear and that CiG, in conjunction with other factors such as the number of vaccines secured, can produce better scenarios.

At the early stages of the pandemic, it was understandable that many countries hedged on various possibilities to secure their access to the supplies of approved vaccines[26]. It seems, however, that this competition is not slowing down. By March 2022, for example, Australia and Switzerland secured the number of vaccines sufficient to cover 990% and 837% of their populations, respectively, while 45% and 42% of the population in these countries had booster shots. This stands in contrast with Chile, which secured approximately half of this number (438%), but 83% of its population received booster shots. The available evidence in terms of CiG in Chile indicates a downward trend in this region in recent years[27]. Similar to other examples, the success of vaccine coverage in this country was linked to the development of a robust immunization system after experiencing the H1N1 flu pandemic in 2009. It should be noted though that during the present pandemic, despite high rates of vaccinations, the country suffered from high rates of casualties due to the emergence of new variants[28].

This study does not propose an optimum number of vaccines need to be secured for a successful vaccination campaign, however it illustrates that more does not necessarily mean better. In line with calls for an equitable global distribution of resources and action plans[29], the present study may help to turn the focus of policymakers towards the need for optimising their use of resources and more investment in public trust, rather than merely acquiring more supplies. Indeed, oversupply might lead to the availability of vaccines being taken for granted and, perhaps, shunned. It is essential to increase the public awareness towards the vitality of immunization programmes in general and the importance of individuals’ active participation which when unwavering could help with increasing the vaccination uptake, on top of the acquisition of resources to meet the population’s needs.

### Study limitations

The present study is based on two snapshots of a dynamic process. Yet, these snapshots were taken at the points when in many countries, particularly in HICs, vaccination campaigns had already been established for some time. Also, the dataset included more evidence from HICs than LMICs. However, as the aim here was to investigate the interaction between trust and securing high volumes of vaccines, the latter variable had more relevance to the condition of HICs. It also should be noted that vaccination coverage depends on a wide range of regional factors, such as infrastructure and coordination, that call for future studies.

## Conclusion

This study finds a fine balance between the volume of resources secured at the procurement point and their extent of uptake at the recipient end, combined with the public’s level of confidence in the national government. The results indicate that, although securing sufficient resource matters, ordering more doses does not necessarily mean better vaccine coverage and efforts should also go to increasing the public’s confidence in government and on raising the public’s awareness of the vitality of immunization programmes in general.

## Data Availability

The data is available under license from Gallup

## Ethics committee approval

Not applicable.

## Patient and Public Involvement

Not applicable.

## Abbreviations

CiG: Confidence in government
GDP: Gross domestic product
HICs: High-income countries
LMICs: Low- and middle-income countries

## Footnotes

1 I also replaced CiG with the Gallup National Institutions Index that indicates public confidence in an array of official organisations including the military, the judicial system, the national government and the honesty of elections that returned comparable results.

