## Appendix A for "The connection between COVID-19 vaccine abundance, vaccination coverage, and public trust in government across the globe"

Figure A1 presents the number of vaccines countries have secured versus rate of the population that has been fully vaccinated per hundred people by continent according to the data from IMF-WHO COVID-19 vaccine supply tracker and Our World in Data as of February 2022[1].

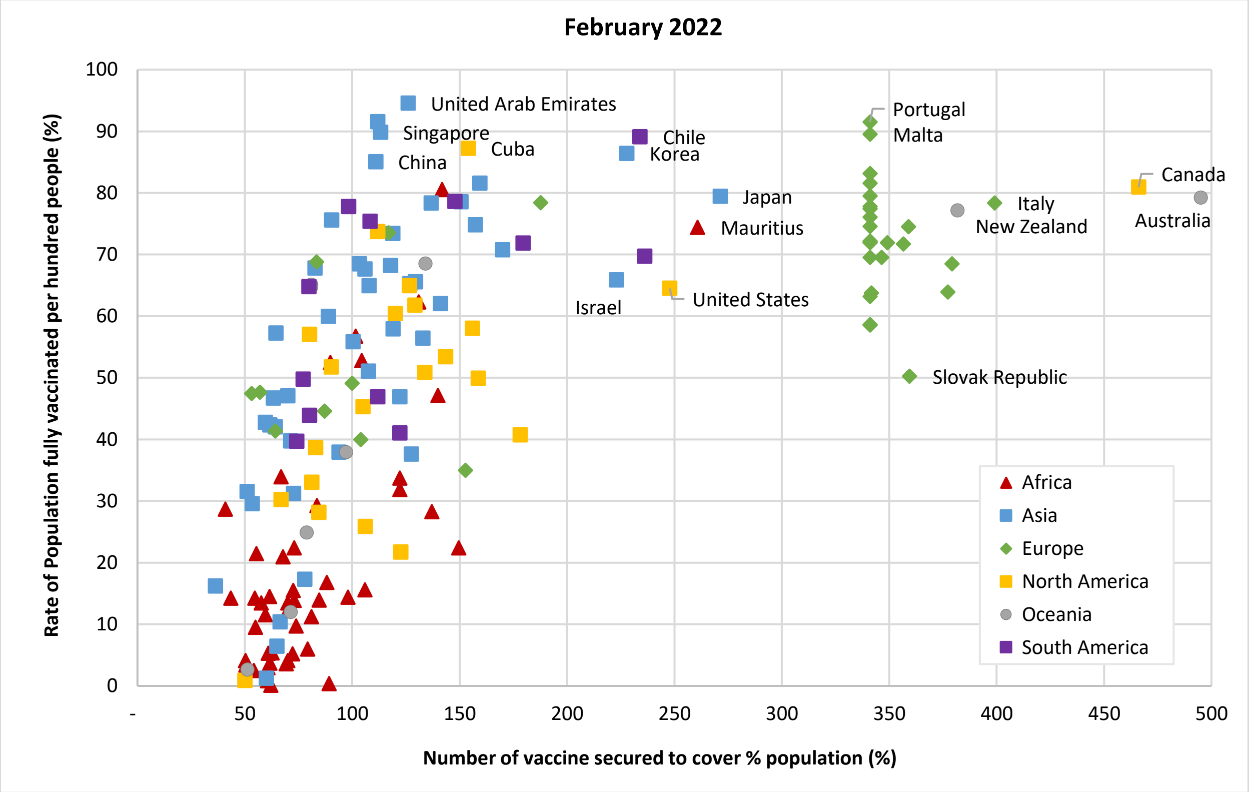


**Figure A1.** Distribution of number of vaccines secured and rate of population vaccinated in (%) as of February 2022[1].

[1] https://ourworldindata.org/covid-vaccinations; https://www.imf.org/en/Topics/imf-and-covid19/IMF-WHO-COVID-19-Vaccine-Tracker.

Table A1 presents the list of countries included in the present study and their characteristics.

**Table A1.** List of countries included in the present study, number of observations (N) and figures for the key variables.

| **Country** | ***N*** | **Fully vaccinated per hundred (%)** | | **Booster (%)** | | **Secured (%)** |
|---|---|---|---|---|---|---|
| **LMICs** | | **Nov** | **Feb** | **Feb** | **Nov** | **Feb** |
| Benin | 1,000 | 5.24 | 14.20 | - | 33.43 | 43.43 |
| Bolivia | 1,004 | 35.09 | 46.89 | 8.62 | 82.18 | 111.91 |
| Bulgaria | 1,005 | 25.70 | 29.58 | 9.82 | 318.59 | 341.05 |
| Colombia | 1,000 | 48.27 | 64.77 | 15.44 | 65.29 | 79.90 |
| Congo B. | 1,000 | 2.28 | 11.23 | - | 61.01 | 81.01 |
| Costa Rica | 1,001 | 62.56 | 73.71 | 20.04 | 127.99 | 111.87 |
| Ghana | 1,000 | 2.65 | 14.40 | 0.32 | 86.50 | 97.99 |
| Kenya | 1,000 | 5.02 | 13.46 | 0.44 | 56.14 | 69.92 |
| Malawi | 1,000 | 3.06 | 4.07 | - | 30.39 | 50.39 |
| Mali | 1,000 | 1.54 | 3.68 | - | 30.00 | 61.68 |
| Mauritius | 1,000 | 68.99 | 74.40 | 32.42 | 252.79 | 260.79 |
| Philippines | 1,000 | 32.75 | 56.42 | 8.81 | 107.27 | 132.90 |
| Romania | 1,009 | 38.85 | 42.06 | - | 318.59 | 341.05 |
| Russia | 2,001 | 38.85 | 49.07 | 8.96 | 70.00 | 100.00 |
| Serbia | 1,000 | 45.46 | 47.43 | 27.01 | 52.49 | 53.08 |
| Ukraine | 1,000 | 26.00 | 34.96 | 1.68 | 84.61 | 152.75 |
| **HICs** | | | | | | |
| Australia | 1,000 | 72.94 | 79.23 | 42.42 | 495.02 | 495.02 |
| Austria | 1,000 | 64.85 | 72.16 | 54.64 | 318.59 | 341.05 |
| Canada | 1,008 | 76.24 | 80.92 | 45.32 | 413.57 | 466.10 |
| Croatia | 1,002 | 47.33 | 54.52 | - | 318.59 | 341.05 |
| Cyprus | 1,019 | 65.07 | 71.88 | 48.60 | 326.65 | 349.11 |
| Czech Rep. | 1,008 | 59.52 | 63.75 | 37.14 | 318.61 | 341.73 |
| Denmark | 1,010 | 76.26 | 81.56 | 62.34 | 318.59 | 341.05 |
| Estonia | 1,024 | 59.46 | 63.17 | 33.08 | 318.59 | 341.05 |
| Finland | 1,005 | 72.70 | 76.03 | 49.36 | 318.59 | 341.05 |
| France | 1,000 | 69.93 | 77.39 | 52.28 | 318.59 | 341.05 |
| Germany | 1,000 | 68.13 | 74.52 | 55.85 | 336.47 | 358.92 |
| Greece | 1,006 | 63.70 | 72.00 | 50.45 | 318.59 | 341.05 |
| Hungary | 1,000 | 60.69 | 63.92 | 39.37 | 354.92 | 377.37 |
| Iceland | 500 | 76.25 | 78.37 | 67.23 | 187.78 | 187.78 |
| Ireland | 1,000 | 76.68 | 79.47 | 56.12 | 318.59 | 341.05 |
| Italy | 1,000 | 73.06 | 78.35 | 61.42 | 376.57 | 399.02 |
| Japan | 1,007 | 77.31 | 79.44 | 15.38 | 271.32 | 271.32 |
| Latvia | 1,017 | 63.85 | 69.49 | 26.29 | 318.59 | 346.43 |

| | | | | | | |
|---|---|---|---|---|---|---|
| Malta | 1,001 | 83.77 | 89.49 | 66.08 | 318.59 | 341.05 |
| Netherlands | 1,000 | 70.12 | 71.91 | 51.81 | 318.59 | 341.05 |
| New Zealand | 1,000 | 70.69 | 77.16 | 43.51 | 333.50 | 381.85 |
| Norway | 1,007 | 70.30 | 73.49 | 52.43 | 77.26 | 116.86 |
| Poland | 1,001 | 54.02 | 58.52 | 29.56 | 318.59 | 341.05 |
| Portugal | 1,002 | 88.38 | 91.49 | 60.05 | 318.59 | 341.05 |
| Slovakia | 1,003 | 46.76 | 50.22 | 29.62 | 337.04 | 359.50 |
| South Korea | 1,001 | 80.26 | 86.38 | 59.92 | 208.35 | 227.84 |
| Spain | 1,000 | 80.49 | 83.12 | 51.07 | 318.59 | 341.05 |
| Sweden | 1,010 | 69.92 | 74.55 | 47.62 | 318.59 | 341.05 |
| Switzerland | 1,000 | 65.55 | 68.48 | 41.07 | 268.49 | 379.21 |
| UK | 1,000 | 68.03 | 64.52 | 28.04 | 273.09 | 247.81 |
| US | 1,005 | 60.18 | 71.70 | 55.79 | 247.81 | 356.66 |
| **Total** | 47,656 | | | | | |

Table A2 presents the complete list of coefficients for running the model full vaccination rate as of November 2021.

Table A2. Full list of coefficients of association between the rate of people fully vaccinated per hundred in Nov. 2021 and the interaction between CiG, and number of vaccines secured for population by thresholds of 200% and 300% as well as other covariates.

| Variables | Number secured and GDP excluded | Number secured excluded | Secured 200% or more | Secured 300% or more |
|---|---|---|---|---|
| | (1) | (2) | (3) | (4) |
| CiG = 1 | 1.89*** | -1.70*** | -2.69*** | -3.61*** |
| | (1.43 - 2.35) | (-2.01 - -1.39) | (-3.26 - -2.12) | (-4.08 - -3.14) |
| Secured 200% or more = 1 | | | 12.33*** | |
| | | | (11.80 - 12.86) | |
| CiG # Secured 200% or more | | | 3.36*** | |
| | | | (2.71 - 4.01) | |
| Gender = Female | -0.23 | 0.14 | 0.15 | 0.10 |
| | (-0.70 - 0.23) | (-0.17 - 0.45) | (-0.13 - 0.44) | (-0.20 - 0.39) |
| Age | 0.36*** | 0.13*** | 0.09*** | 0.10*** |
| | (0.34 - 0.37) | (0.12 - 0.14) | (0.08 - 0.09) | (0.09 - 0.11) |
| Employed (any type of employment) | -0.17 | -0.40** | -0.13 | -0.17 |
| | (-0.70 - 0.35) | (-0.74 - -0.05) | (-0.45 - 0.19) | (-0.50 - 0.16) |
| Per Capita Income Quintiles† = Second 20% | -2.05*** | -0.68** | -0.46* | -0.53** |
| | (-2.81 - -1.28) | (-1.20 - -0.15) | (-0.94 - 0.03) | (-1.02 - -0.03) |
| Per Capita Income Quintiles = Middle 20% | -3.44*** | -1.10*** | -0.80*** | -0.92*** |
| | (-4.20 - -2.68) | (-1.61 - -0.58) | (-1.28 - -0.32) | (-1.41 - -0.43) |
| Per Capita Income Quintiles = Fourth 20% | -4.50*** | -1.46*** | -1.10*** | -1.25*** |
| | (-5.25 - -3.74) | (-1.97 - -0.96) | (-1.57 - -0.63) | (-1.73 - -0.77) |
| Per Capita Income Quintiles = Richest 20% | -6.50*** | -2.04*** | -1.58*** | -1.77*** |
| | (-7.26 - -5.75) | (-2.54 - -1.54) | (-2.04 - -1.11) | (-2.24 - -1.30) |
| Married / Domestic partner = 1 (yes) | -2.86*** | -0.71*** | -0.71*** | -0.78*** |
| | (-3.34 - -2.39) | (-1.02 - -0.39) | (-1.00 - -0.41) | (-1.09 - -0.48) |
| Health Problem = 1 (yes) | -4.56*** | -2.41*** | -1.82*** | -2.15*** |
| | (-5.15 - -3.96) | (-2.79 - -2.03) | (-2.18 - -1.47) | (-2.51 - -1.79) |
| Internet Access = 1 (yes) | 24.96*** | 10.46*** | 8.72*** | 8.72*** |
| | (24.12 - 25.81) | (9.89 - 11.04) | (8.17 - 9.27) | (8.16 - 9.28) |
| Born in Country = 1 (yes) | -7.00*** | -2.00*** | -2.07*** | -1.63*** |
| | (-7.75 - -6.26) | (-2.56 - -1.44) | (-2.56 - -1.58) | (-2.13 - -1.14) |
| Education Level†† = Secondary, 3-year tertiary secondary education and some education beyond secondary education. | 5.51*** | 0.05 | -0.77*** | -0.84*** |
| | (4.72 - 6.29) | (-0.48 - 0.57) | (-1.26 - -0.28) | (-1.33 - -0.35) |

| | | | | |
|---|---|---|---|---|
| Education Level = Completed four years of education beyond high school and/or received a 4 - year college degree. | 8.04*** | -0.95*** | -1.33*** | -1.21*** |
| | (7.22 - 8.86) | (-1.52 - -0.37) | (-1.87 - -0.80) | (-1.75 - -0.68) |
| Rural/Urban[†††] = A small town or village | 4.42*** | 3.05*** | 1.97*** | 2.29*** |
| | (3.70 - 5.14) | (2.59 - 3.51) | (1.54 - 2.40) | (1.86 - 2.72) |
| Rural/Urban = A large city | 4.10*** | 3.94*** | 2.73*** | 3.04*** |
| | (3.35 - 4.86) | (3.47 - 4.42) | (2.28 - 3.17) | (2.59 - 3.49) |
| Rural/Urban = A suburb of a large city | 9.25*** | 5.02*** | 4.11*** | 4.79*** |
| | (8.42 - 10.09) | (4.47 - 5.57) | (3.59 - 4.62) | (4.28 - 5.31) |
| Population Density | 0.03*** | 0.02*** | 0.02*** | 0.02*** |
| | (0.02 - 0.03) | (0.02 - 0.02) | (0.02 - 0.02) | (0.02 - 0.02) |
| GDP per Capita | | 0.00*** | 0.00*** | 0.00*** |
| | | (0.00 - 0.00) | (0.00 - 0.00) | (0.00 - 0.00) |
| Secured 300% or more = 1 | | | | 6.84*** |
| | | | | (6.42 - 7.26) |
| CiG # Secured 300% or more | | | | 4.50*** |
| | | | | (3.91 - 5.10) |
| Constant | 16.02*** | 8.99*** | 11.44*** | 10.93*** |
| | (14.54 - 17.50) | (7.97 - 10.01) | (10.44 - 12.43) | (9.94 - 11.92) |
| Observations | 46,819 | 46,819 | 46,819 | 46,819 |
| R-squared | 0.35 | 0.70 | 0.74 | 0.73 |

Robust 95% CI in parentheses; *** p<0.01, ** p<0.05, * p<0.1.

[†] Ref. Poorest 20%; [††]Ref. Completed elementary education or less; [†††]Ref. A rural area or on a farm

Tables A3 and A4 presents the complete list of coefficients for running the model full vaccination rate and also rate of booster shots as of February 2022.

**Table A3.** Full list of coefficients of association between the rate of people fully vaccinated per hundred in Feb. 2022 and the interaction between CiG, and number of vaccines secured for population by thresholds of 200% and 300% as well as other covariates.

| Variables | Number secured and GDP excluded<br>(1) | Number secured excluded<br>(2) | Secured 200% or more<br>(3) | Secured 300% or more<br>(4) |
|---|---|---|---|---|
| CiG = 1 | 1.94*** | -1.21*** | -2.65*** | -2.63*** |
| | (1.50 - 2.38) | (-1.53 - -0.89) | (-3.32 - -1.99) | (-3.19 - -2.06) |
| Secured 200% or more = 1 | | | 8.71*** | |
| | | | (8.13 - 9.29) | |
| CiG # Secured 200% or more | | | 3.55*** | |
| | | | (2.81 - 4.29) | |
| Gender = Female | -0.17 | 0.16 | 0.18 | 0.15 |
| | (-0.61 - 0.27) | (-0.16 - 0.48) | (-0.13 - 0.49) | (-0.17 - 0.47) |
| Age | 0.33*** | 0.13*** | 0.09*** | 0.11*** |
| | (0.31 - 0.34) | (0.12 - 0.14) | (0.08 - 0.10) | (0.10 - 0.12) |
| Employed (any type of employment) | -0.28 | -0.47** | -0.26 | -0.23 |
| | (-0.77 - 0.22) | (-0.84 - -0.11) | (-0.61 - 0.09) | (-0.59 - 0.13) |
| Per Capita Income Quintiles† = Second 20% | -1.95*** | -0.74*** | -0.57** | -0.66** |
| | (-2.69 - -1.21) | (-1.29 - -0.19) | (-1.10 - -0.04) | (-1.20 - -0.12) |
| Per Capita Income Quintiles = Middle 20% | -3.29*** | -1.23*** | -1.00*** | -1.14*** |
| | (-4.02 - -2.56) | (-1.77 - -0.69) | (-1.52 - -0.48) | (-1.67 - -0.61) |
| Per Capita Income Quintiles = Fourth 20% | -4.32*** | -1.66*** | -1.38*** | -1.57*** |
| | (-5.05 - -3.60) | (-2.19 - -1.13) | (-1.90 - -0.87) | (-2.09 - -1.05) |
| Per Capita Income Quintiles = Richest 20% | -6.29*** | -2.36*** | -2.02*** | -2.25*** |
| | (-7.01 - -5.57) | (-2.89 - -1.84) | (-2.52 - -1.52) | (-2.76 - -1.74) |
| Married / Domestic partner = 1 (yes) | -2.88*** | -0.99*** | -0.99*** | -0.99*** |
| | (-3.34 - -2.43) | (-1.32 - -0.65) | (-1.31 - -0.66) | (-1.32 - -0.67) |
| Health Problem = 1 (yes) | -4.62*** | -2.73*** | -2.28*** | -2.52*** |
| | (-5.19 - -4.04) | (-3.13 - -2.33) | (-2.67 - -1.89) | (-2.91 - -2.13) |
| Internet Access = 1 (yes) | 23.73*** | 10.99*** | 9.68*** | 9.97*** |
| | (22.90 - 24.57) | (10.36 - 11.62) | (9.05 - 10.30) | (9.34 - 10.60) |
| Born in Country = 1 (yes) | -6.57*** | -2.18*** | -2.22*** | -1.88*** |
| | (-7.28 - -5.87) | (-2.74 - -1.62) | (-2.72 - -1.71) | (-2.40 - -1.35) |
| Education Level†† = Secondary, 3-year tertiary secondary education and some education beyond secondary education. | 5.77*** | 0.97*** | 0.33 | 0.53* |
| | (5.02 - 6.52) | (0.42 - 1.52) | (-0.21 - 0.86) | (-0.01 - 1.07) |

| | | | | |
|---|---|---|---|---|
| Education Level = Completed four years of education beyond high school and/or received a 4 - year college degree. | 7.91*** | 0.01 | -0.32 | -0.17 |
| | (7.13 - 8.69) | (-0.58 - 0.61) | (-0.89 - 0.26) | (-0.75 - 0.41) |
| Rural/Urban[†††] = A small town or village | 3.95*** | 2.75*** | 1.94*** | 2.35*** |
| | (3.26 - 4.65) | (2.26 - 3.24) | (1.46 - 2.41) | (1.87 - 2.83) |
| Rural/Urban = A large city | 3.97*** | 3.82*** | 2.91*** | 3.48*** |
| | (3.24 - 4.69) | (3.32 - 4.33) | (2.42 - 3.40) | (2.98 - 3.97) |
| Rural/Urban = A suburb of a large city | 9.03*** | 5.31*** | 4.61*** | 5.39*** |
| | (8.23 - 9.83) | (4.73 - 5.89) | (4.05 - 5.17) | (4.83 - 5.95) |
| Population Density | 0.03*** | 0.02*** | 0.02*** | 0.02*** |
| | (0.02 - 0.03) | (0.02 - 0.02) | (0.02 - 0.02) | (0.02 - 0.02) |
| GDP per Capita | | 0.00*** | 0.00*** | 0.00*** |
| | | (0.00 - 0.00) | (0.00 - 0.00) | (0.00 - 0.00) |
| Secured 300% or more = 1 | | | | 4.51*** |
| | | | | (4.04 - 4.99) |
| CiG # Secured 300% or more | | | | 2.95*** |
| | | | | (2.28 - 3.62) |
| Constant | 24.26*** | 18.09*** | 20.29*** | 19.25*** |
| | (22.81 - 25.71) | (17.00 - 19.18) | (19.18 - 21.40) | (18.14 - 20.36) |
| Observations | 46,819 | 46,819 | 46,819 | 46,819 |
| R-squared | 0.35 | 0.65 | 0.67 | 0.66 |

Robust 95% CI in parentheses; *** p<0.01, ** p<0.05, * p<0.1.
[†] Ref. Poorest 20%; [††]Ref. Completed elementary education or less; [†††]Ref. A rural area or on a farm

**Table A4.** Full list of coefficients of association between the rate of booster shots per hundred in Feb. 2022 and the interaction between CiG, and number of vaccines secured for population by thresholds of 200% and 300% as well as other covariates.

| Variables | Number secured and GDP excluded<br>(1) | Number secured excluded<br>(2) | Secured 200% or more<br>(3) | Secured 300% or more<br>(4) |
|---|---|---|---|---|
| CiG = 1 | 4.10***<br>(3.64 - 4.55) | 0.44***<br>(0.12 - 0.77) | 1.65***<br>(1.18 - 2.12) | 2.11***<br>(1.63 - 2.58) |
| Secured 200% or more = 1 | | | 13.37***<br>(12.86 - 13.89) | |
| CiG # Secured 200% or more | | | -0.22<br>(-0.80 - 0.37) | |
| Gender = Female | -0.67***<br>(-1.13 - -0.21) | -0.15<br>(-0.47 - 0.17) | -0.14<br>(-0.43 - 0.16) | -0.20<br>(-0.47 - 0.08) |
| Age | 0.20***<br>(0.18 - 0.21) | 0.04***<br>(0.03 - 0.05) | 0.00<br>(-0.01 - 0.01) | -0.00<br>(-0.01 - 0.01) |
| Employed (any type of employment) | -0.82***<br>(-1.34 - -0.30) | -1.03***<br>(-1.39 - -0.68) | -0.87***<br>(-1.20 - -0.55) | -0.50***<br>(-0.80 - -0.19) |
| Per Capita Income Quintiles† = Second 20% | -0.94**<br>(-1.71 - -0.17) | -0.02<br>(-0.56 - 0.52) | 0.17<br>(-0.32 - 0.67) | 0.12<br>(-0.34 - 0.58) |
| Per Capita Income Quintiles = Middle 20% | -1.62***<br>(-2.37 - -0.87) | -0.07<br>(-0.60 - 0.46) | 0.21<br>(-0.28 - 0.69) | 0.18<br>(-0.27 - 0.62) |
| Per Capita Income Quintiles = Fourth 20% | -1.80***<br>(-2.55 - -1.04) | 0.10<br>(-0.43 - 0.62) | 0.42*<br>(-0.07 - 0.90) | 0.32<br>(-0.13 - 0.76) |
| Per Capita Income Quintiles = Richest 20% | -2.59***<br>(-3.34 - -1.84) | 0.22<br>(-0.31 - 0.75) | 0.62**<br>(0.13 - 1.11) | 0.49**<br>(0.04 - 0.94) |
| Married / Domestic partner = 1 (yes) | -1.05***<br>(-1.53 - -0.57) | 0.29*<br>(-0.04 - 0.62) | 0.31**<br>(0.00 - 0.61) | 0.29**<br>(0.01 - 0.57) |
| Health Problem = 1 (yes) | -2.52***<br>(-3.12 - -1.93) | -1.52***<br>(-1.92 - -1.12) | -0.95***<br>(-1.32 - -0.58) | -0.96***<br>(-1.30 - -0.62) |
| Internet Access = 1 (yes) | 14.31***<br>(13.44 - 15.19) | 4.48***<br>(3.89 - 5.06) | 3.17***<br>(2.63 - 3.72) | 1.98***<br>(1.48 - 2.47) |
| Born in Country = 1 (yes) | -4.79***<br>(-5.49 - -4.08) | -0.87***<br>(-1.43 - -0.32) | -0.99***<br>(-1.47 - -0.51) | -0.09<br>(-0.52 - 0.34) |
| Education Level†† = Secondary, 3-year tertiary secondary education and some education beyond secondary education. | 0.78*<br>(-0.02 - 1.58) | -2.45***<br>(-2.99 - -1.90) | -3.09***<br>(-3.61 - -2.57) | -3.29***<br>(-3.80 - -2.79) |
| Education Level = Completed four years of education beyond high school and/or received a 4 - year college degree. | 1.38*** | -4.46*** | -4.64*** | -4.51*** |

| | | | | |
|---|---|---|---|---|
| | (0.54 - 2.23) | (-5.06 - -3.87) | (-5.21 - -4.07) | (-5.06 - -3.96) |
| Rural/Urban[†††] = A small town or village | 2.77*** | 1.88*** | 0.59*** | 0.54*** |
| | (2.04 - 3.51) | (1.42 - 2.34) | (0.18 - 1.00) | (0.16 - 0.91) |
| Rural/Urban = A large city | 2.91*** | 2.90*** | 1.47*** | 1.72*** |
| | (2.15 - 3.67) | (2.42 - 3.38) | (1.04 - 1.91) | (1.32 - 2.13) |
| Rural/Urban = A suburb of a large city | 5.22*** | 2.34*** | 1.13*** | 2.35*** |
| | (4.41 - 6.04) | (1.79 - 2.89) | (0.63 - 1.63) | (1.90 - 2.80) |
| Population Density | 0.02*** | 0.02*** | 0.02*** | 0.02*** |
| | (0.02 - 0.02) | (0.02 - 0.02) | (0.01 - 0.02) | (0.02 - 0.02) |
| GDP per Capita | | 0.00*** | 0.00*** | 0.00*** |
| | | (0.00 - 0.00) | (0.00 - 0.00) | (0.00 - 0.00) |
| Secured 300% or more = 1 | | | | 16.16*** |
| | | | | (15.69 - 16.63) |
| CiG # Secured 300% or more | | | | -1.80*** |
| | | | | (-2.37 - -1.23) |
| Constant | 13.65*** | 1.24** | 2.27*** | 1.34*** |
| | (12.12 - 15.19) | (0.19 - 2.28) | (1.29 - 3.25) | (0.44 - 2.24) |
| Observations | 40,973 | 40,973 | 40,973 | 40,973 |
| R-squared | 0.17 | 0.59 | 0.65 | 0.70 |

Robust 95% CI in parentheses; *** p<0.01, ** p<0.05, * p<0.1.
[†] Ref. Poorest 20%; [††]Ref. Completed elementary education or less; [†††]Ref. A rural area or on a farm

Figure A2 presents the rate of booster shots per hundred people as of February 2022 and the number of vaccines secured across countries and their continents.

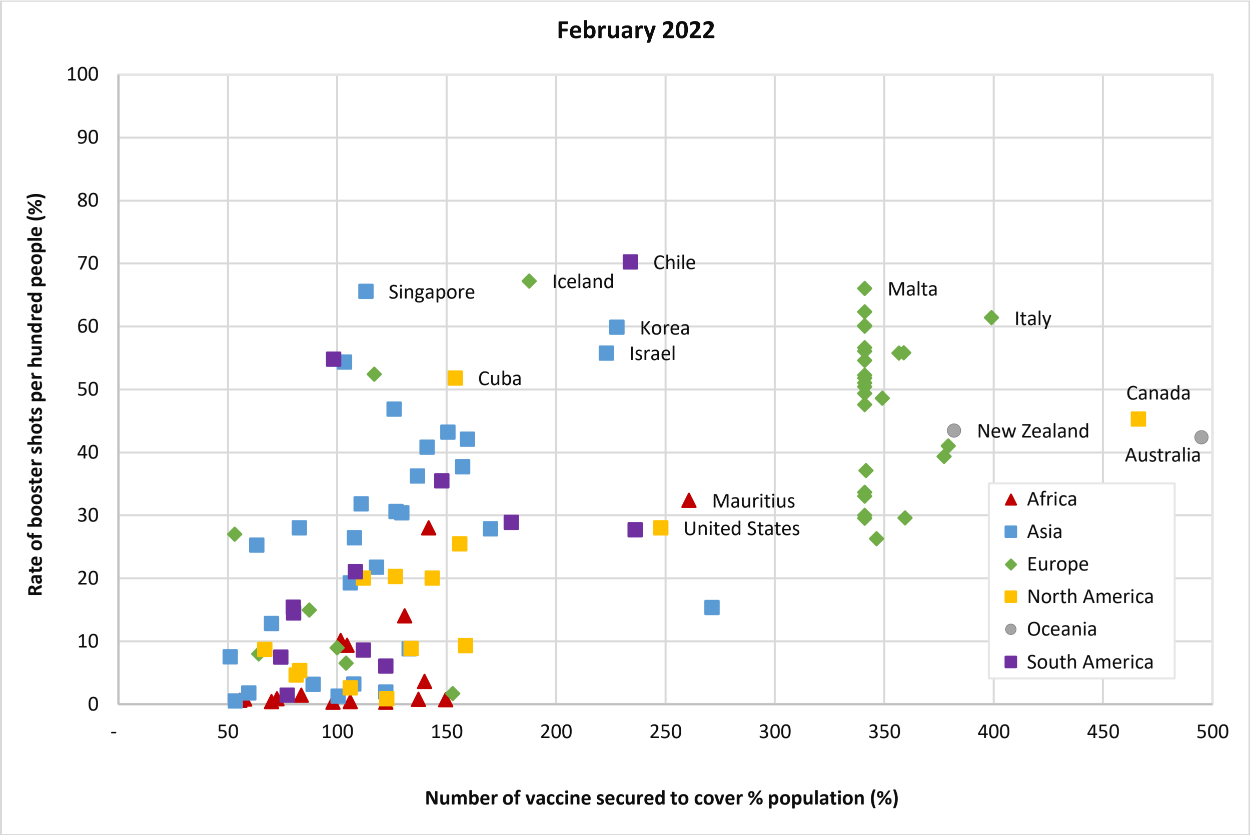


Figure 2. Distribution of number of vaccines secured and rate of population who received booster shots per hundred in (%) as of February 2022.

Table A5 show the results for robustness checks for repeating the analysis for full vaccination rates as of February 2022 where in the former, two countries (Canada and Australia) who had outstanding number of vaccines secured were removed from the data, and in the latter, number of nurses per 10,000 people is also included in the model. As it can be seen, the main results hold and remain comparable to those presented in Table A3.

**Table A5.** Robustness checks for the association between the rate of people fully vaccinated per hundred in Feb. 2022 and the interaction between CiG, and number of vaccines secured for population by thresholds of 200% and 300% as well as other covariates.

| | Removing Canada and Australia as outliers | | Inclusion of ratio of nurses per population per 10,000 people | |
|---|---|---|---|---|
| Variables | Secured 200% or more (1) | Secured 300% or more (2) | Secured 200% or more (3) | Secured 300% or more (4) |
| CiG = 1 | -2.64*** | -2.63*** | -2.56*** | -2.56*** |
| | (-3.31 - -1.97) | (-3.19 - -2.06) | (-3.23 - -1.90) | (-3.12 - -1.99) |
| Secured 200% or more = 1 | 8.42*** | | 8.03*** | |
| | (7.84 - 9.00) | | (7.44 - 8.61) | |
| CiG # Secured 200% or more | 3.47*** | | 3.67*** | |
| | (2.73 - 4.22) | | (2.93 - 4.40) | |
| Gender = Female | 0.21 | 0.19 | 0.18 | 0.16 |
| | (-0.11 - 0.53) | (-0.14 - 0.52) | (-0.13 - 0.49) | (-0.16 - 0.48) |
| Age | 0.10*** | 0.11*** | 0.10*** | 0.11*** |
| | (0.09 - 0.11) | (0.10 - 0.12) | (0.09 - 0.11) | (0.10 - 0.12) |
| Employed (any type of employment) | -0.33* | -0.31 | -0.25 | -0.23 |
| | (-0.70 - 0.03) | (-0.68 - 0.06) | (-0.60 - 0.10) | (-0.59 - 0.13) |
| Per Capita Income Quintiles† = Second 20% | -0.56** | -0.65** | -0.58** | -0.67** |
| | (-1.11 - -0.01) | (-1.21 - -0.09) | (-1.11 - -0.05) | (-1.21 - -0.13) |
| Per Capita Income Quintiles = Middle 20% | -1.03*** | -1.17*** | -1.02*** | -1.15*** |
| | (-1.57 - -0.49) | (-1.72 - -0.62) | (-1.53 - -0.50) | (-1.68 - -0.62) |
| Per Capita Income Quintiles = Fourth 20% | -1.43*** | -1.61*** | -1.39*** | -1.57*** |
| | (-1.96 - -0.90) | (-2.15 - -1.07) | (-1.90 - -0.88) | (-2.09 - -1.05) |
| Per Capita Income Quintiles = Richest 20% | -2.05*** | -2.29*** | -2.03*** | -2.24*** |
| | (-2.57 - -1.53) | (-2.82 - -1.75) | (-2.53 - -1.53) | (-2.75 - -1.74) |
| Married / Domestic partner = 1 (yes) | -1.00*** | -1.00*** | -1.01*** | -1.02*** |
| | (-1.33 - -0.66) | (-1.35 - -0.66) | (-1.33 - -0.69) | (-1.35 - -0.70) |
| Health Problem = 1 (yes) | -2.38*** | -2.63*** | -2.24*** | -2.46*** |
| | (-2.78 - -1.98) | (-3.03 - -2.22) | (-2.62 - -1.85) | (-2.85 - -2.07) |
| Internet Access = 1 (yes) | 10.01*** | 10.33*** | 9.93*** | 10.30*** |
| | (9.37 - 10.64) | (9.69 - 10.97) | (9.31 - 10.56) | (9.67 - 10.93) |
| Born in Country = 1 (yes) | -2.14*** | -1.77*** | -2.08*** | -1.75*** |

| | | | | |
|---|---|---|---|---|
| | (-2.70 - -1.59) | (-2.34 - -1.20) | (-2.58 - -1.57) | (-2.26 - -1.23) |
| Education Level[††] = Secondary, 3-year tertiary secondary education and some education beyond secondary education. | 0.16 | 0.36 | 0.25 | 0.43 |
| | (-0.39 - 0.70) | (-0.19 - 0.91) | (-0.29 - 0.78) | (-0.11 - 0.97) |
| Education Level = Completed four years of education beyond high school and/or received a 4 - year college degree. | -0.49* | -0.36 | -0.38 | -0.25 |
| | (-1.07 - 0.09) | (-0.95 - 0.24) | (-0.95 - 0.19) | (-0.83 - 0.33) |
| Rural/Urban[†††] = A small town or village | 2.05*** | 2.47*** | 1.84*** | 2.22*** |
| | (1.56 - 2.54) | (1.98 - 2.96) | (1.36 - 2.31) | (1.74 - 2.70) |
| Rural/Urban = A large city | 2.97*** | 3.54*** | 2.74*** | 3.26*** |
| | (2.47 - 3.47) | (3.04 - 4.05) | (2.25 - 3.23) | (2.77 - 3.75) |
| Rural/Urban = A suburb of a large city | 4.45*** | 5.22*** | 4.55*** | 5.26*** |
| | (3.85 - 5.04) | (4.62 - 5.82) | (4.00 - 5.11) | (4.70 - 5.82) |
| Population Density | 0.02*** | 0.02*** | 0.02*** | 0.02*** |
| | (0.02 - 0.02) | (0.02 - 0.02) | (0.02 - 0.02) | (0.02 - 0.02) |
| GDP per Capita | 0.00*** | 0.00*** | 0.00*** | 0.00*** |
| | (0.00 - 0.00) | (0.00 - 0.00) | (0.00 - 0.00) | (0.00 - 0.00) |
| Nurses per population per 10,000 | | | -0.05*** | -0.06*** |
| | | | (-0.06 - -0.04) | (-0.07 - -0.05) |
| Secured 300% or more = 1 | | 4.20*** | | 3.72*** |
| | | (3.72 - 4.68) | | (3.23 - 4.20) |
| CiG # Secured 300% or more | | 2.85*** | | 3.22*** |
| | | (2.16 - 3.53) | | (2.55 - 3.89) |
| Constant | 20.06*** | 18.97*** | 20.30*** | 19.34*** |
| | (18.91 - 21.22) | (17.82 - 20.12) | (19.19 - 21.40) | (18.24 - 20.45) |
| Observations | 44,872 | 44,872 | 46,819 | 46,819 |
| R-squared | 0.66 | 0.65 | 0.67 | 0.66 |

Robust 95% CI in parentheses; *** p<0.01, ** p<0.05, * p<0.1.

[†] Ref. Poorest 20%; [††]Ref. Completed elementary education or less; [†††]Ref. A rural area or on a farm
